# Assessment of Adverse Childhood Experiences in the South Bronx on Risk of Developing Chronic Disease as Adults

**DOI:** 10.1101/2023.05.12.23289819

**Authors:** Alexander Njoroge, Masood A Shariff, Hira W Khan, Victor Gordillo, Brian Eclarinal, Jose Vargas, Mohammad Faiz, Moiz Kasubhai, Tranice Jackson

**Author notes:** Corresponding author: Masood A Shariff MD Department of Medicine NYC HHC Lincoln, 234 E 149^TH^ ST, Bronx, NY 10541 USA. **Presentation:** Quality of Care and Outcomes Research Scientific Sessions in Reston, Virginia held on May 13–14, 2022. Conflict of Interest statement: The authors have no affiliations.

## Abstract

**Background:** Adverse Childhood Experiences (ACE) have a negative impact on health outcomes. Using a cross-sectional study design, our objective was to identify the prevalence of ACEs among residents of the South Bronx and the relationship between such childhood stressors and prevalence of both chronic disease and modifiable high-risk behavior in adulthood.

**Methods:** We recruited patients from a hospital-based adult primary care clinic in the metropolitan area of South Bronx. A cross-sectional survey was conducted between September 2017 and January 2018, using a modified ACE Questionnaire, that included nine ACE categories (Physical Abuse, Sexual Abuse, Household Substance Abuse, Separation from Parents, Incarcerated Household Member, Parental Separation/Divorce, and Bullying), and in addition to questions on demographics, high-risk behavior, and diagnosis of chronic disease. Our primary objective was to gather incidence of ACEs organized by domains. Secondary objectives were to demonstrate any expected increase (as Odds Ratio) in Chronic Disease or maladaptive social habits when compared to patients with no ACEs within the cohort.

**Results:** A total of 454 patients completed the survey. The average age was 53.1±14.2 years and females were 49% of the sample. Hispanics were 61% followed by Blacks at 34%. Participants reported high-risk behavior in 24%, had a high prevalence of chronic illness (82%) and ACE events at 70%. We found a significant relationship between ACE events and having a chronic disease diagnosis and engagement in high-risk behavior with higher odds of reporting chronic illnesses among participants with exposure to childhood stressors (OR 1.26, 95% confidence interval 1.1-1.5, p=0.002). Of the nine ACE categories many were independently associated with one or more chronic diseases in adulthood.

**Conclusion:** According to our survey data, ACE events in our patient population were more prevalent (30% with 4 or more exposures), higher than the proposed average of 1 out of 6 Americans with 4 or more exposures nationally according to the national statistics. These childhood stressors appear to have a strong association with development of high-risk behavior and chronic illnesses.

## Introduction

Adverse Childhood Experiences (ACEs) have been shown to contribute significantly to negative adult health behaviors and health outcomes. In prospective cohort studies, people exposed to multiple childhood traumatic stressors were at increased risk of premature death compared to those without ACEs (1). Another investigation has linked ACE to early death and yet others have linked ACE with depressive disorders (2), diagnoses of cancer (3,4), Chronic Obstructive Pulmonary Disease (COPD) (5,6), premature mortality (7,8), self-reported sleep disturbances (9), cardiovascular health (10,11), and high-risk behaviors (12,13). Studies have also investigated the association between adverse childhood events and neurological damage, with significant effects on social behavior, affect and memory (14). Additionally, strong relationships have been demonstrated between ACEs and risky sexual behaviors, substance abuse and aggression (12–16).

Ongoing research continues to highlight the burden of ACEs and its impact on adults, specifically between the associations of ACE and the impact they play in the high-risk behavior and outcomes of these patients with chronic disease. In the United States, an estimated 65 to 87% of adults have experienced at least one ACE in their lifetime (3). ACE, alongside Social Determinants of Health (SDOH) and environmental factors, all have been proven to influence a person’s health (17).

The South Bronx is the congressional district with the lowest socioeconomic standing per capita in the country, with high rates of substance abuse and other risk factors for health and chronic diseases (18–22). To our best knowledge, ACE events have not been measured in this community. Thus, our findings will help examine the relationship to chronic disease and high-risk behavior among adults attending a large hospital based primary care clinic who present with more than one chronic illness.

## Methods

The Institutional Review Board approved the study (IRB#16-009) and data collection was conducted in a single-center outpatient adult primary care clinic in the South Bronx, NY. Eligibility criteria included age over 18 years and electronic medical record verification of care being received at the study center for at least one year. The recruitment was a consecutive sampling of patients that presented to the clinic for primary care appointment. They were called a day prior to the visit to introduce the study and gauge their interest in willingness to participate in answering the survey questionnaire prior to or following their clinic appointment. Research staff conducted face-to-face interviews and administered a modified ACE questionnaire (discussed below). Cognizant of the sensitive nature of the interview, all research staff underwent training on interview techniques and followed a protocoled script to discuss with prospective patients, including engagement, confidentiality, the research question, and the goals of the study. The questionnaire was pre-tested among medical students and outpatient clinical staff for clarity and timing. The study and its goals were reviewed again with the patients presented for their primary care appointment, consented and interviewed in an enclosed space with a suitable level of privacy. Upon completion, survey data were entered into an electronic database and de-identified. More than fifty percent of the South Bronx patient population is largely of Hispanic descent/Spanish speakers. Our research staff consisted of certified native Spanish speakers who engaged these patients and conducted their surveys.

### Questionnaire Design

According to the American Association of Pediatrics (AAP), there are ten recognized adverse childhood events (23,25). These stressors were originally formulated using information from the first ACE study (1) and include: *Emotional Abuse; Mother Treated Violently; Physical Abuse; Household Substance Abuse; Sexual Abuse; Household Mental Illnesses; Emotional Neglect; Parental Separation or Divorce; Physical Neglect; and Incarcerated Household Member.* Our survey design was developed institutionally, with input from the Psychiatric and Pediatric departments, with emphasis on two ACE definitions/domains-*Abuse* and *Household Dysfunction-* given the prevalence in our patient population. We modified wording in the ACE questions about sexual abuse, verbal abuse and mother treated violently; these changes were introduced from other formats of the ACE survey found to be more concise while preserving integrity of main themes. Adjustments also included the addition of four categories, *parental separation or divorce*, *bullying in school*, *separation from parents,* and *living with foster parents,* as these were not part of the standard ACE questionnaire. These categories were included following the review of the AAP publications on childhood stressors (24). The AAP has emphasized all four categories as significant causes of childhood distress, especially in the early years (25,26). The complete ACE study tool is shown in Supplemental Table 1.

The study questionnaire was modified from the original ACE study (1), Adverse Childhood Experience Questionnaire for Adults and other formats of the survey (27) were utilized referring to the subject’s life before 18 years of age. There were three categories within the Abuse definition: A-Verbal abuse, B-Physical abuse, C-Sexual abuse. Eight categories within the Household Dysfunction definition: D-Household substance abuse, E-Household mental illness, F-Family member treated violently, G-Separation from parents, H-Incarcerated family member, I-Parental separation or divorce, J-Bulling in school, and K-Foster parents. Figure 1 illustrates the distribution of ACE categories.

**Figure 1.**
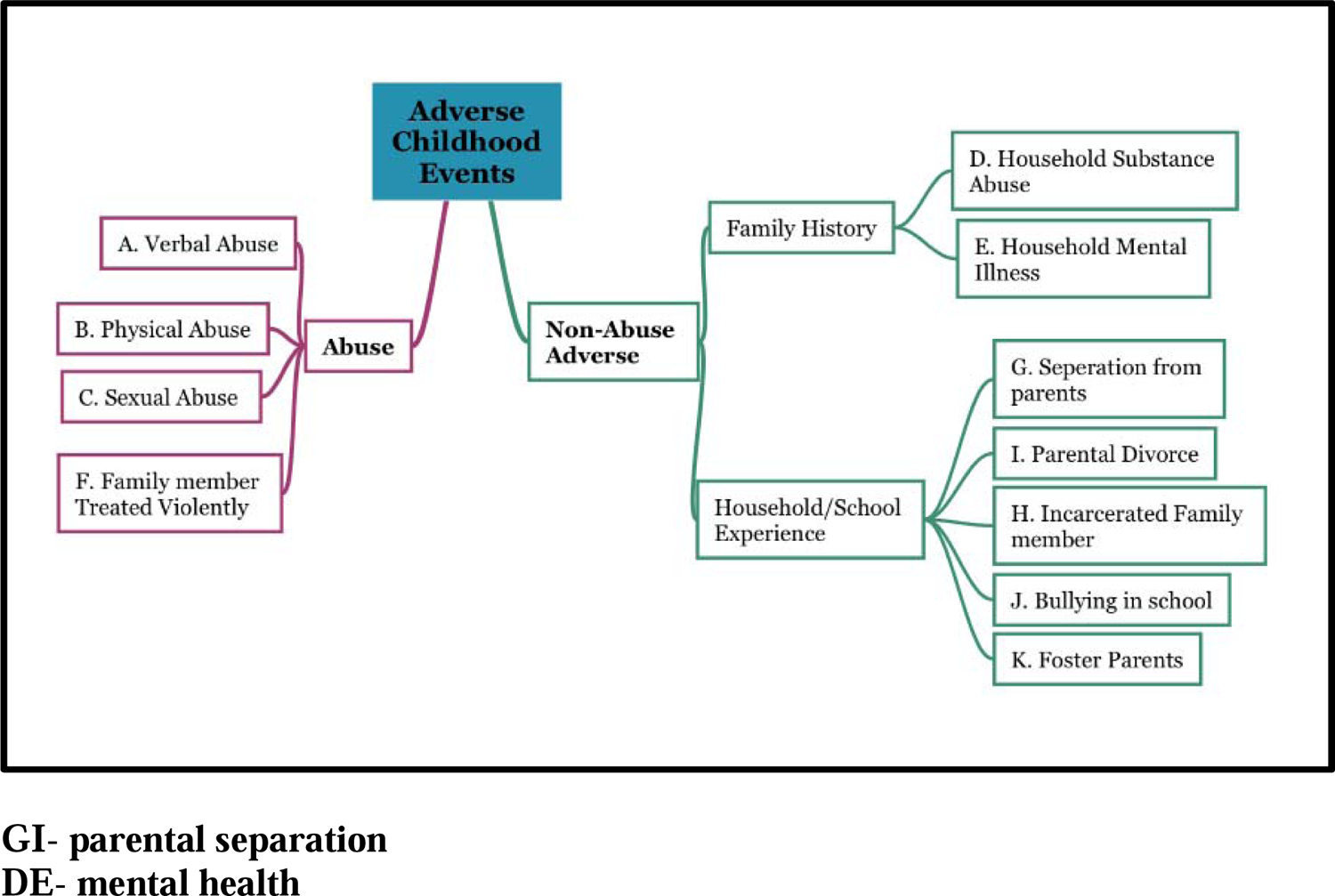
Distribution of ACE definitions and categories.

Studies have been published using various combinations of the original questionnaire. There are four questions within our abuse definition that inquired about verbal abuse (Category A), physical abuse (Category B) and sexual abuse (Category C). Category B (physical abuse) is subdivided into two questions. Under household dysfunction, family members were treated violently, household substance abuse, household mental illness, parental separation or divorce, and incarcerated household members were inquired. Category D (Household substance abuse) is subdivided into two questions. Category F (Family member treated violently) is subdivided into four questions, for a total of thirteen household dysfunction questions. All responses were binary, and participants were marked as exposed if they responded “yes” to any of the subdivided category questions. The measure of exposure, or ACE Score, is the sum of positive categories; and the scores ranged from 0 (unexposed) to 11 (exposed to all categories).

### Risky Behavior and Chronic Disease Assessment

We added questions to the prior survey about participants’ subjective health (i.e. how would you describe your health?) and medical diagnoses (ie. has a doctor, nurse or other healthcare provider ever told you that you have asthma?). Chronic illnesses were assessed by collecting history of Hypertension, Dysglycemia/Diabetes, Hyperlipidemia, Heart Attack Event, Angina/Coronary Heart Disease (CHD), Stroke, Asthma, Chronic Obstructive Pulmonary Disease (COPD), Congestive Heart Failure (CHF), Hepatitis, and Poor Mental Health/Antipsychotic Use (in the past 30 days).

We also included three questions assessing high risk behavior such as smoking status, alcohol and substance or drug abuse. High-risk behavior was screened with three pertinent questions on smoking history (daily, somedays, quit, or none), daily alcohol use (one or more drinks as per CDC definition [30]), and substance abuse (marijuana, heroin, cocaine, methamphetamine, or prescription drugs).

### Demographics Assessed

Demographic information was assessed through the use of eleven questions on age, gender, race, primary language, place of birth, years in the United States (US), insurance type, education, marital status, and employment status/description. The questionnaire is attached in the Supplemental section.

### Statistical Analysis

Our sample size was 454, and we used a confidence interval of 95% and alpha error of 0.05 to evaluate associations between childhood stressors and health outcomes, childhood stressors and risky behavior. We used multiple logistic regression to assess significant relationships between various chronic illnesses, total ACE score, specific ACE categories and other covariates. The odds ratio (OR) for the associations was calculated by logistic regression using SPSS Statistics for Windows Version 15 (SPSS Inc, Chicago, Illinois, USA). The 95% confidence interval (CI) and P-value for each OR were tabulated. P<.05 was deemed significant. Individual logistic regression models for each of the eleven health variables listed were used to measure the exposure-response relationship of ACE scores with health outcomes.

## Results

### Demographics

Overall patients screened on a consecutive basis were 610 in the ambulatory clinic setting and 454 patients were consented. The basic demographic breakdown is demonstrated in Table 1. Average age of the participants was 53.1±14.2 years, with a female prevalence of 49%. A majority of the participants were Hispanic (61%), followed by Black (34%), then other (5%). Other demographics of note for our patient population are Asians and some Whites. The most common primary language was Spanish (56%), and a majority of the foreign-born (76%) had lived in the US for more than 10 years (86%). The insurance under “other” represented some combination of private/hospital coverage with Medicare and/or Medicaid (64%). Furthermore, 12% had Medicaid alone, and 13% reported no coverage at all. About 51% of the participants stated they had not finished high school (US equivalent). Marital status was distributed with “Never Married” (36%), “Married” (35%) being the most common response followed by “Divorced” (12%) and “Widow/Widower” (9%). Overall, 44% of participants were employed and described jobs in varied sectors, and of those working nearly half reported more than one job (49%). Participants categorized their job type as “semi-skilled” (16%), “service” (13%), and “skilled” (12%).

**Table 1:**
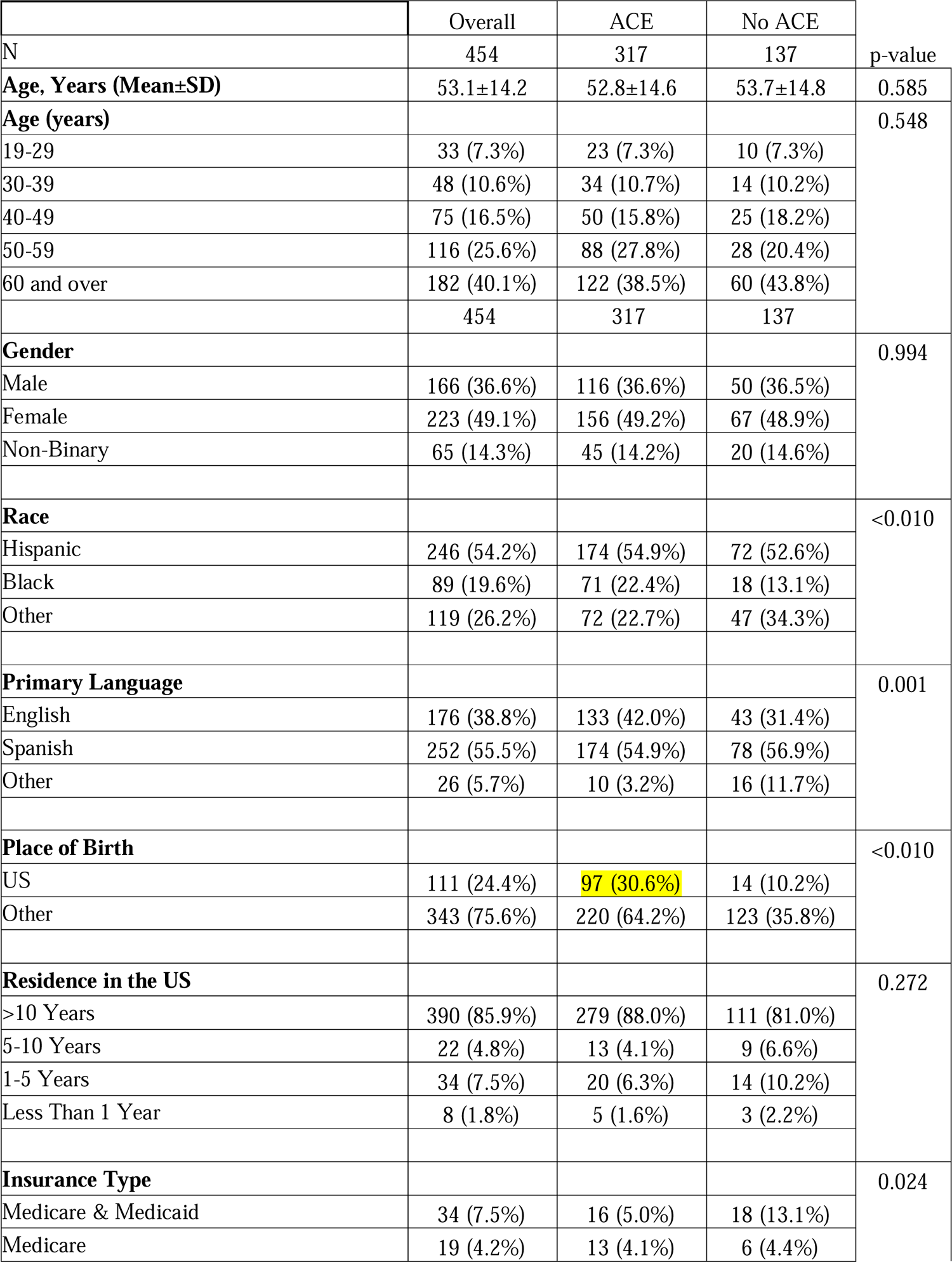
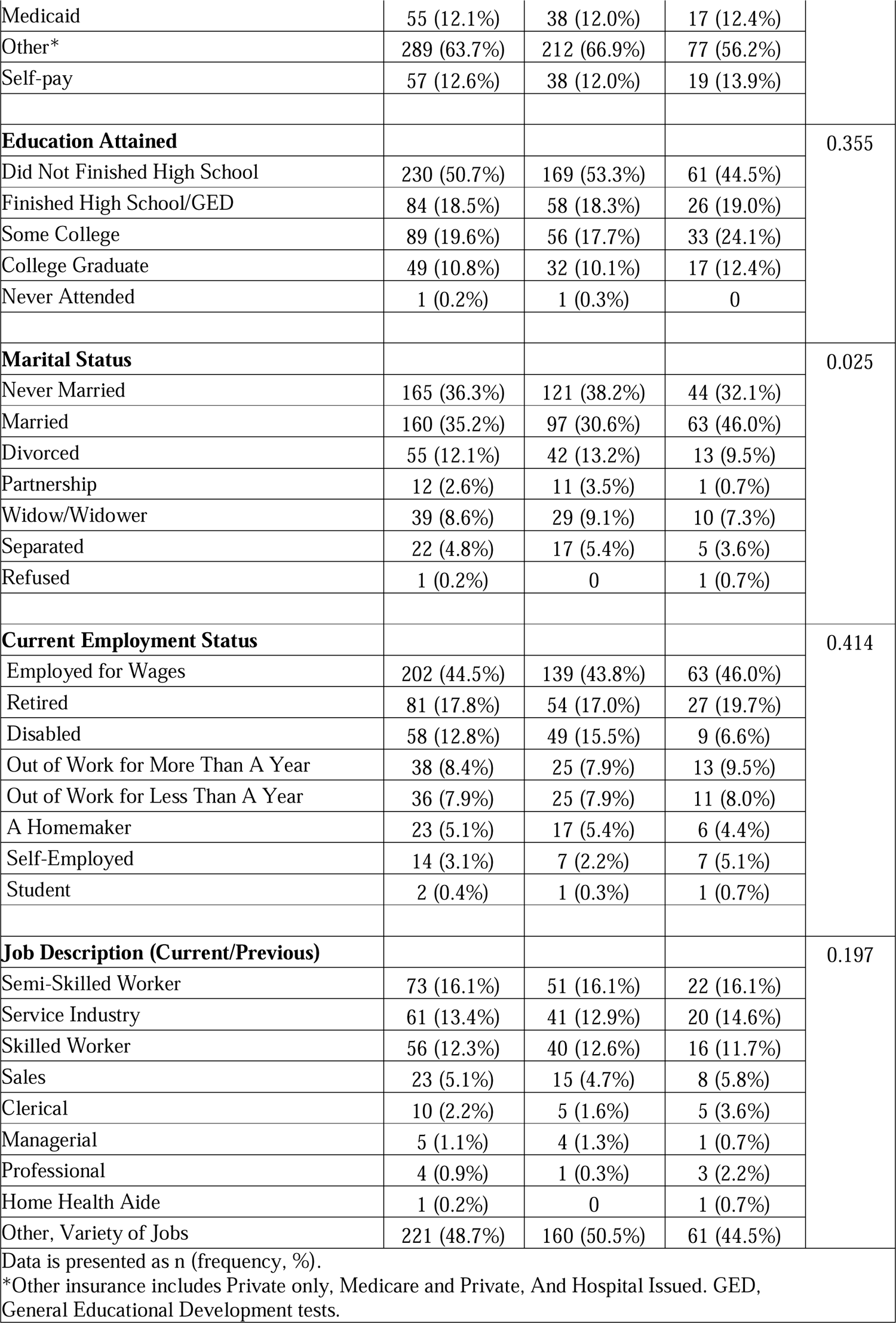
Participant characteristics

### Social Habits

Table 2 presents the participant’s social history, overall health perception, high risk behavior and chronic disease status. A majority of the patients stated they never smoked (76%), 9% had quit smoking, and 16% were current smokers. Daily alcohol consumption was reported in 9% of patients, whereas daily substance abuse/drug use was at 3%. General health self-assessment was “Fair” at 47% and “Good” at 31%. Chronic diseases reported from highest to lowest were: hypertension (59%), hyperlipidemia (48%), dysglycemia/diabetes (43%), asthma (22%), angina/CHD (10%), followed by heart attack, stroke, COPD, hepatitis and CHF. Poor mental health and/or antipsychotic use was 11% for the overall group.

**Table 2:** Participants current overall general health, high risk behavior and chronic disease status

| Table 2: Participants current social, overall general health and chronic disease status | Overall | ACE | No ACE |  |
| --- | --- | --- | --- | --- |
| N/n | 454 | 314 | 140 | p-value |
| **In General, Your Health is:** |  |  |  | 0.394 |
| Excellent | 39 (8.6%) | 27 (8.6%) | 12 (8.6%) |  |
| Very Good | 14 (3.1%) | 10 (3.2%) | 4 (2.9%) |  |
| Good | 139 (30.6%) | 87 (27.7%) | 52 (37.1%) |  |
| Fair | 215 (47.4%) | 153 (48.7%) | 62 (44.3%) |  |
| Poor | 26 (5.7%) | 19 (6.1%) | 7 (5.0%) |  |
| Bad | 12 (2.6%) | 11 (3.5%) | 1 (0.7%) |  |
| Not Sure | 6 (1.3%) | 4 (1.3%) | 2 (1.4%) |  |
| Don't Know | 3 (0.7%) | 3 (1.0%) | 0 |  |
| **Chronic Disease** |  |  |  |  |
| Hypertension | 266 (58.6%) | 182 (58.0%) | 84 (60.0%) | 0.766 |
| Hyperlipidemia | 219 (48.2%) | 157 (50.0%) | 62 (44.3%) | 0.400 |
| Diabetes | 195 (43.0%) | 141 (44.9%) | 54 (38.6%) | 0.317 |
| Asthma | 101 (22.2%) | 73 (23.2%) | 28 (20.0%) | 0.271 |
| Angina/CHD | 46 (10.1%) | 33 (10.5%) | 13 (9.3%) | 0.968 |
| Heart Attack Event | 32 (7.0%) | 19 (6.1%) | 13 (9.3%) | 0.182 |
| Stroke | 27 (5.9%) | 20 (6.4%) | 7 (5.0%) | 0.620 |
| COPD | 22 (4.8%) | 19 (6.1%) | 3 (2.1%) | 0.083 |
| Hepatitis | 23 (5.1%) | 14 (4.5%) | 9 (6.4%) | 0.621 |
| CHF | 15 (3.3%) | 9 (2.9%) | 6 (4.3%) | 0.399 |
| **High Risk Behavior** |  |  |  |  |
| Alcohol Dependence | 41 (9.0%) | 34 (10.8%) | 7 (5.0%) | 0.082 |
| Mental Health | 49 (10.8%) | 45 (14.3%) | 4 (2.9%) | 0.001 |
| Substance Abuse | 15 (3.3%) | 14 (4.5%) | 1 (0.7%) | 0.016 |
| **Smoking Status** |  |  |  | 0.024* |
| Never Smoked | 343 (75.6%) | 226 (72.0%) | 117 (83.6%) |  |
| Quit | 40 (8.8%) | 31 (9.9%) | 9 (6.4%) |  |
| Smoking Status | 71 (15.6%) | 57 (18.0%) | 14 (10.2%) |  |
| Smoker-Some Days | 33 (7.3%) | 21 (6.7%) | 12 (8.6%) |  |
| Smoker-Every Day | 38 (8.4%) | 36 (11.5%) | 2 (1.4%) |  |
| Data is presented as n (frequency, %). CHD, Coronary artery disease; CHF, congestive heart failure; COPD, chronic obstructive pulmonary disease. Smoking variables were combined to Never smoked, Quit, and Smoker. |  |  |  |  |
| * |  |  |  |  |

### ACE Data

Figure 2 presents all the ACE components surveyed in the study. The commonest of ACE experienced were as follows: 59% of participants said they experienced “parent separation/divorce” (Category I) as children; followed by “separated from parent” (Category G) (40%) and “family members treated violently” (Category F) (19%). Participants who did not report any ACE events were 43% (n=137), while the remainder 57% (n=317) had a median event of one ACE and it ranged from 1 to 11 with one event being the majority at 21% overall. The ACE events were summed as total events per participant and the average was 2.1±2.4 events overall.

**Figure 2:**
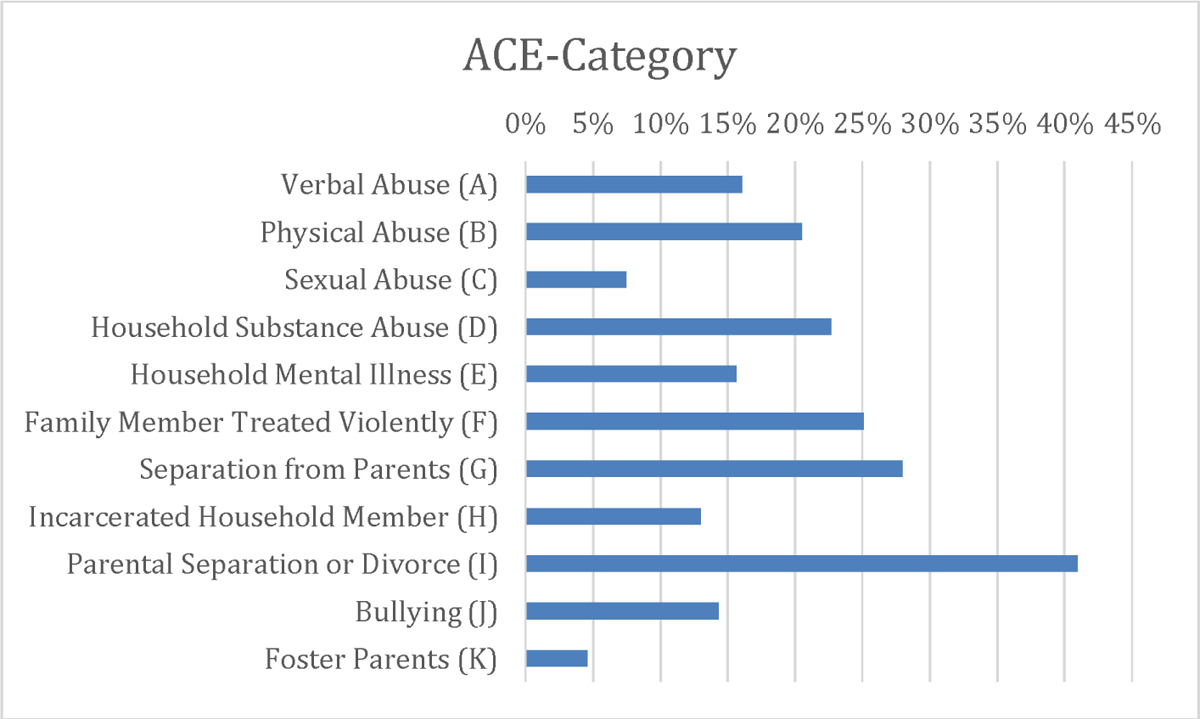

Subgroup comparison between the “No ACE” versus “One or More ACE” (Table 1) revealed there was no difference in relative risk between men and women. Distribution by race similarly demonstrated no difference between subgroups. Participants born in the US had a higher incidence of ACE versus those born outside the US (87% vs. 64%). More than half (53%) of those who did not finish high school reported one or more ACE. Those married faced one or more ACE (31%), while “never married” (38% one or more ACE vs. 32% no ACE) and “divorced” (13% vs. 9%) were higher in the events subgroup compared to no ACE, respectively. Employment status and types of employment were comparable between the groups without significance.

Subgroup comparison (Table 2), the overall health assessment indicated a difference of 10% for self-reported “Good” health between the ACE (28%) and no ACE groups (36%). Smoking every day was also significantly higher in the events group at 12% versus 1% in the no events group (p=0.024). Participants with an ACE event had a higher incidence of both reported mental health (14% vs. 3%, p<0.01) conditions than those with no exposure and substance abuse (5% vs. 1%, p=0.016).

### ODDS RATIOS: By ACE specific association with chronic disease

The findings demonstrated a high prevalence of both chronic illnesses (80%) and at least one ACE (70%) at baseline within the surveyed population. Incidence of the most common chronic diseases reported-Hypertension (59%), hyperlipidemia (48%), and diabetes (43%). Between no ACE and at least one ACE, the burden of chronic disease was the same. Certain ACE categories were correlated with having a larger number of chronic diseases (hypertension / hyperlipidemia / diabetes), they include: Sexual Abuse (OR 21.53, 4.14, and 4.65, respectively); Household Substance Abuse (OR 2.82, 1.80, and 2.05, respectively); Mental Illness (OR 2.95, 2.29, and 2.00, respectively); and Parent’s Separated or Divorce (OR 3.83, 2.53, and 2.18, respectively) (p<0.01). Additionally, Physical Abuse (B) predicted having two chronic diseases (OR 2.52 CI 1.3-5.0, p<0.01). Finally, participants with one or more chronic diseases significantly correlated with Separation from Parents (G) (OR 2.94, CI 1.66-5.22, p<0.01), Incarcerated Household Member (H) (OR 2.61, CI 1.12-6.08, p<0.05) and Bullying (J) (OR 4.84, CI 1.89-12.38, p<0.01). Table 3 lists odds ratios between chronic disease and ACE Categories.

**Table 3:** Odds ratio for number of chronic disease and ACE Categories

| ACE Categories | With One Chronic Disease |  |  | With Two Chronic Disease |  |  | With Three Chronic Diseases |  |  |
| --- | --- | --- | --- | --- | --- | --- | --- | --- | --- |
|  | OR | 95% C.I. |  | OR | 95% C.I. |  | OR | 95% C.I. |  |
| A - Verbal Abuse | 0.695 | 0.28 | 1.73 | 0.59 | 0.28 | 1.27 | 0.55 | 0.251 | 1.22 |
| B - Physical Abuse | 2.03 | 0.894 | 4.62 | 2.52** | 1.27 | 5.00 | 2.11* | 1.06 | 4.21 |
| C - Sexual Abuse | 21.53** | 2.62 | 176.89 | 4.14** | 1.47 | 11.68 | 4.65** | 1.87 | 11.54 |
| D - Household Substance Abuse | 2.82** | 1.46 | 5.44 | 1.80* | 1.05 | 3.08 | 2.05** | 1.19 | 3.51 |
| E - Household Mental Illness | 2.95* | 1.26 | 6.90 | 2.29* | 1.21 | 4.35 | 2.01* | 1.09 | 3.70 |
| F - Treated violently by family member | 1.43 | 0.71 | 2.87 | 0.86 | 0.48 | 1.53 | 1.00 | 0.55 | 1.80 |
| G - Separation from parents | 2.94** | 1.66 | 5.22 | 1.80* | 1.1 | 2.96 | 1.08 | 0.63 | 1.86 |
| H - Incarcerated Household member | 2.61* | 1.12 | 6.08 | 1.68 | 0.87 | 3.26 | 1.20 | 0.61 | 2.34 |
| I - Parental separation/divorce | 3.83** | 2.31 | 6.37 | 2.53** | 1.6 | 4.00 | 2.18** | 1.33 | 3.59 |
| J - Bullying | 4.84** | 1.89 | 12.38 | 1.70 | 0.86 | 3.33 | 2.00* | 1.04 | 3.88 |
| K - Foster parent | 0.737 | 0.15 | 3.61 | 0.70 | 0.23 | 2.16 | 0.23* | 0.07 | 0.87 |
| Constant | 0 |  |  | 0.001 |  |  | 0.005 |  |  |

Significant \*p<0.05 and \*\*p<.01

The odds of having anyone of the 11 chronic diseases were higher in respondents with one or more ACE when compared to individuals with no ACE. The ACE score (number of ACE) also appeared to have a significant relationship with chronic disease (OR 1.26, CI 1.1-1.5, p<0.01); however, there was no association between ACE score of greater than four and less than four with chronic disease. “ACE increases risk of CD but the risk did not increase with number of ACE events” Further analysis involved comparing individual chronic illnesses with ACE scores and ACE categories. Most individual ACE categories showed significant relationships with eight of the eleven chronic illnesses to varying degrees, with highest odds ratio relating to hypertension, hyperlipidemia, dysglycemia, CHD, stroke, asthma, COPD and history of mental health/antipsychotic use. High-risk behaviors (alcohol use disorder, substance use disorder and smoking history) also correlated individually with ACE categories significantly (Table 4).

**Table 4:** Incidence of ACE Categories with Chronic Disease and Adverse Social Habits (Number of patients with X condition who had Y category of ACE)

|  | ACE Categories |  |  |  |  |  |  |  |  |  |  |
| --- | --- | --- | --- | --- | --- | --- | --- | --- | --- | --- | --- |
|  | Verbal Abuse (A) | Physical Abuse (B) | C-Sexual Abuse (C) | Household Substance Abuse (D) | Household Mental Illness (E) | Treated violently by family member (F) | Separation from parents (G) | Incarcerated Household member (H) | Parental separation /divorce (I) | Bullying (J) | Foster parent (K) |
| Hypertension | 41 | 60 | 29** | 59 | 51* | 70 | 74 | 36 | 111 | 44 | 11 |
| Hyperlipidemia | 36 | 54* | 24** | 52 | 40 | 56 | 61 | 32 | 89 | 32 | 12 |
| Diabetes | 32 | 45* | 21* | 47 | 35 | 49 | 53 | 30 | 75 | 33 | 10 |
| Heart attack event | 6 | 7 | 3 | 8 | 8 | 11 | 10 | 4 | 14 | 5 | 1 |
| Angina/CHD | 7 | 14 | 7* | 13 | 11 | 14 | 12 | 10 | 19 | 13** | 5* |
| Stroke | 6 | 7 | 5* | 8 | 8* | 7 | 9 | 2 | 16* | 6 | 1 |
| Asthma | 23* | 32** | 10 | 29 | 28** | 31 | 33 | 15 | 45 | 26** | 6 |
| COPD | 8** | 11** | 2 | 9* | 6 | 8 | 9 | 6* | 11 | 7* | 1 |
| CHF | 1 | 3 | 2 | 3 | 2 | 5 | 1 | 3 | 5 | 3 | 0 |
| Hepatitis | 3 | 3 | 2 | 10* | 3 | 6 | 5 | 4 | 8 | 3 | 0 |
| Mental Health (> 14 days for the last 30 days) | 15** | 20** | 7 | 21** | 19** | 26** | 16 | 7 | 28* | 17** | 3 |
| Alcohol- Daily Use | 16** | 19** | 7* | 17** | 18** | 20** | 16 | 9 | 19 | 13** | 3 |
| Substance Abuse History | 9** | 9** | 4* | 8* | 8** | 7 | 10* | 7** | 12* | 8** | 5** |
| Smoking History | 27** | 32* | 10 | 38** | 24* | 34 | 40* | 23** | 58** | 21 | 6 |

Significant \*p<0.05 and \*\*p<0.010. CHD, Coronary artery disease; CHF, congestive heart failure; COPD, chronic obstructive pulmonary disease.

## Discussion

To our knowledge, this is the first study assessing ACE in a population in the South Bronx. In our sample of 454 patients at an urban primary care clinic, 70% had experienced at least one ACE event and, of those, 80% had chronic disease. Having an ACE event/score of at least one predicted a current diagnosis of chronic medical disease. This was likely driven by particular subcategories of ACE that showed an impact on chronic disease, mental health, and substance abuse. We found that the people giving an affirmative response to Parent Divorce/Separation was a common reported category in our sample size, followed by Separated from Parent, Family Member Treated Violently, Household Substance Abuse, and Physical Abuse having a more than 20% response rate by the participants for those categories.

### Mental Health and Chronic Disease

There was a significant correlation with chronic illnesses and mental health diseases in all the ACE categories, which has also been shown in other studies with ACE Score of 4 plus (29,30). Among our study participants who reported one or more high-risk behaviors, a majority also reported having at least one ACE event, consistent with the established relationship between childhood stressors and high-risk behaviors in other studies (12,13,28).

### ACE vs No ACE

In comparing ACE and no ACE for high-risk behaviors, we found significant dominance of behaviors such as smoking, alcohol, and substance abuse in the ACE group. Further, this showed a significant association with the ACE A through K categories.

### Results Contrary to existing literature

Our results demonstrated no gender difference for risk of ACE, differing from the increased risk for females demonstrated in most existing literature (37). The majority of the respondents who reported one or more ACE were of Hispanic descent, compared to Black and others. Cultural perceptions between different ethnicities may influence the recognition and reporting of childhood stressors. Ye et al. (38) suggested that there are variations in cultural influences and certain parenting practices, such as disciplinary actions, which may or may not be perceived as abuse. Furthermore, the reporting of ACEs by individuals of various ethnic or cultural backgrounds may be influenced by their shame or fear of social embarrassment (38). In our findings, the relationship between ACEs and age groups showed a high number of ACEs reported by individuals older than 45 years, contrary to other ACE studies and surveys, which linked higher ACE scores to younger adults (33).

The chronic diseases in our study were well distributed between the more than one ACE and no ACE groups. Hypertension, hyperlipidemia, diabetes, and asthma were on top of the list, as Hispanics and Blacks are predisposed to diabetes and hypertension (35–37). Although we did not find any significance in participants who had one or more ACE events compared to the ones who had none for chronic disease, there was correlation between having one or more ACE and recent history of mental health or use of psychiatric medications for anxiety or depression, which was statistically significant. The participants who experienced ACE were diagnosed with a mental health condition and had taken prescribed medication through a primary care provider (rather than with a psychiatrist). This correlation has been found in other studies where individuals who experienced ACE events were diagnosed with a mental health condition and sought help (38–40). Mental health impacts socioeconomic factors, influencing the capacity to maintain employment, aggressive behavior, marital issues, and educational attainment (41–43).

### Findings Consistent with Prior Studies

Consistent with previous findings, ACEs prevalent among the residents of South Bronx is associated with certain poor health outcomes as they have been shown to do in other communities. Having one or more ACE is likely found among residents of Hispanic descent and older age groups. Sexual abuse, family history of mental health, and bullying were associated with hypertension, hyperlipidemia, and dysglycemia with a higher odds ratio. No significant associations were demonstrated between ACEs and diabetes, heart attack, stroke, heart failure, and hepatitis. The Bronx community signifies a low socioeconomic population that is made up of immigrants who have come to the US often for seeking better economic opportunity. It follows then that they are likely to present with a burden of ACEs from their country of origin. ACE events are more prevalent in low- and middle-income countries owing to limited resources and less social health care (44–46). Thus, the idea of being assimilated into the US culture and its correlation with chronic disease actually could have an underlying basis in immigrants with their “ACE burden” that they have experienced. Thus, highlighting the added effort the healthcare community needs to put forth while addressing their medical needs.

## Conclusion and Future Steps

That ACE is prevalent and linked to a higher probability of having chronic disease. Our findings suggest that the health care system could be wise to incorporate an ACE questionnaire into their current SDOH screenings in primary care and develop services for patients to address the mental health impact of ACE. One study found that enabling services (such as coordinating care, improving health education, transportation and access to food, shelter and health care benefits) found an association with percentage increase in health center visits, higher probability of getting routine checkup (i.e. flu shots) and patient satisfaction, concluding with confirmed impact of systematic delivery in health services by reducing access barriers (50). ACE and SDOH have been shown to intersect, and as a result, have been determined to influence health conditions and behaviors into adulthood, thus it becomes imperative to address both SDOH and ACEs. In line with several other studies that used a similar ACE questionnaire, our study also found that chronic diseases in adulthood are often linked to ACE exposure. As a result of this correlation, it becomes important for physicians to implement processes to systematically address social needs as part of clinical care, especially conditions that potentially have the greatest influence on a patient’s health. These processes could include social services, education, one’s physical environment, and nutrition.

While there are many ways for healthcare providers to help their patients address their underlying social conditions, providers should also begin by systematically screening patients for health-related social needs. This initial screening and discussion with the patient can ultimately impact their health by determining early on their needs and current socio-economic situation. In addition, by working alongside community organizations, providers can help patients who may have difficulty accessing health care by facilitating their capability to acquire housing, food, transportation, and/or other social services. Finally, providers have more influence when it comes to health policy and are in a unique position to alter a state and federal government’s ability to prioritize improving health services and facilities for all its residents within its jurisdiction. Through proactive collaboration with community organizations, health care policy changes and changers and patient education, it becomes possible to potentially disentangle the intersect that exists between ACE and SDOH.

### Limitations

There are limitations encountered in this study. First, our findings only represent the South Bronx residents who receive healthcare at a single center. Therefore, residents of the community who were affiliated with other institutions were not adequately represented. Second, the investigation and subsequent results, are centered around a modified study tool. Adaptations made to questions on *sexual abuse*, *verbal abuse,* and *mother treated violently* from the original ACE survey included shortened sentence phrasing and rewording, mainly to accommodate translation into various languages encountered in the South Bronx. The modifications, however, were not subject to scrutinized task force validation as reflected in prior published surveys (47–49). Thirdly, the survey inquired about childhood upbringing; and therefore, recollection or recall bias may affect the reporting of ACEs. Further, we did find an impact between ACE and mental illness; however, our study did not resolve specific psychiatric illnesses such as anxiety or depression. A history of “poor mental health or antipsychotic drug use,” which does not give a precise idea of the mental health issues the participants were facing. Finally, our sample size was adequate based on adequate power to conclude associations with chronic diseases significantly.

However, possibly due to sampling bias, the limited responses of other chronic illnesses such as stroke or heart attack could have been curtailed by recruiting patients from the morning, evening, and weekend clinic sessions. ACE has not been evaluated in the South Bronx region to our knowledge, thus we hope to contribute this information with our research to the overall scientific community, and anchor our findings with continued interrogation of ACE associations with other comorbidities such as mental health.

## Data Availability

All data collected has been analyzed and tabulated into the manuscript. Upon request, the Data can be shared after approval from our institution's ethics committee and data agreement.

## Acknowledgements

We would like to thank the study enrollees for their participation and contribution to the research. The authors would like to specifically acknowledge Paul J. Fields, Heather Mavronicolas, Ina Sunita Tamaldeo, and Cynthia Lee for their assistance. A special thanks to Maria Carlos, MPH and Lara Rabiee, JD, MHS for their input in reviewing the work for authenticity.

## Author Statements

This research did not receive any specific grant from funding agencies in the public, commercial or not-for-profit sectors.

## Supplementary Material

Table: Modified ACE Questionnaire

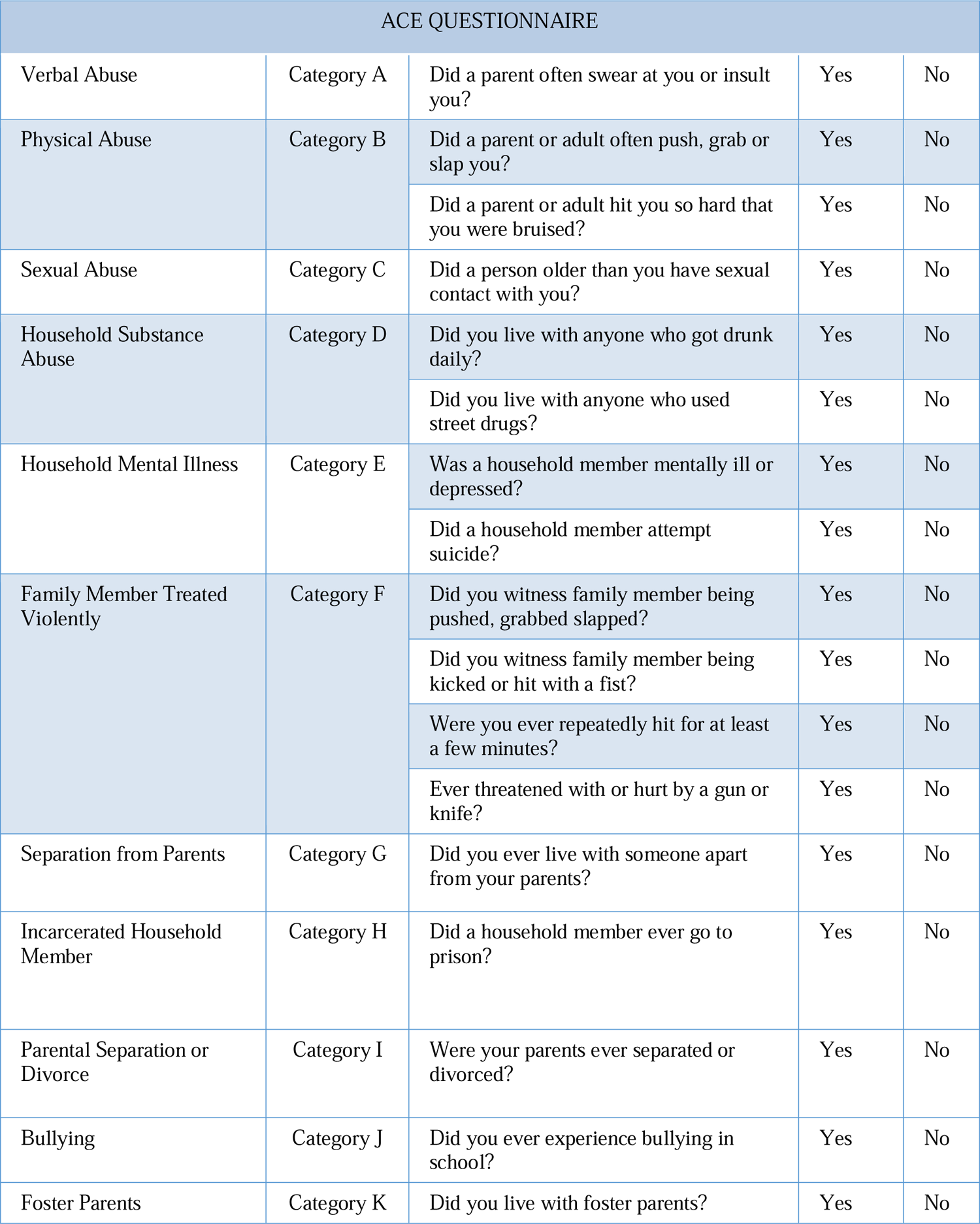

## Notes

### Competing Interest Statement

The authors have declared no competing interest.

### Clinical Trial

The study was conducted as a pilot intervention to assess Adverse Childhood Experiences in the community of south Bronx, Ethics committee approval was received (IRB#16-009).

### Funding Statement

No external funding was received to conduct this study.

### Author Declarations

Ethics committee/IRB of Lincoln Medical Center Office of IRB gave ethical approval for this work

